# Small RNA sequencing reveals snoRNAs and piRNA-019825 as novel players in diabetic kidney disease

**DOI:** 10.1101/2022.05.23.22275440

**Authors:** LM ’t Hart, JA de Klerk, GA Bouland, JHD Peerlings, M.T. Blom, SJ Cramer, R Bijkerk, JWJ Beulens, RC Slieker

## Abstract

**Introduction:** Micro- and macrovascular complications are common among persons with type 2 diabetes. Recently there has been growing interest to investigate the potential of circulating small non-coding RNAs (sncRNAs) as contributors to the development of diabetic complications. In this study we investigate to what extent circulating sncRNAs levels associate with prevalent diabetic kidney disease (DKD) in persons with type 2 diabetes.

**Methods:** Plasma sncRNAs levels were determined using small RNA-seq, allowing detection of miRNAs, snoRNAs, piRNAs, tRNA-fragments and various other sncRNA classes. We tested for differentially expressed sncRNAs in persons with type 2 diabetes, with DKD (n=69) or without DKD (n=405). In secondary analyses, we also tested the association with eGFR, albuminuria (UACR) and the plasma proteome.

**Results:** In total seven sncRNAs were negatively associated with prevalent DKD (all P_FDR_≤0.05). Including one microRNA (miR-143-5p), five snoRNAs (U8, SNORD118, SNORD24, SNORD107, SNORD87) and a piRNA (piR-019825|DQ597218). Proteomic analyses showed that the seven sncRNAs, and especially the piRNA piR-019825, were associated with plasma levels of 24 proteins of which several have known associations with kidney function including TNF sR-I (TNFRFS1A), DAN (NBL1) and cystatin C (CST3).

**Conclusion:** We have identified novel small non-coding RNAs, primarily from classes other than microRNAs, that are associated with diabetic kidney disease. Our results shows that the involvement of small non-coding RNAs in DKD goes beyond the already known microRNAs and also involves other classes of sncRNA, in particular snoRNAs and the piRNA piR-019825, that have never been studied before in relation to kidney function.

## Introduction

Diabetic kidney disease (DKD) is one of the most common complications in type 2 diabetes. Nonetheless, while some develop complications early in the disease, others will never. The mechanisms underlying DKD are not fully understood though [1].

Circulating small non-coding RNAs (sncRNA) are mostly contained in extracellular vesicles or are bound to RNA-binding proteins and lipoproteins and originate from various tissues including tissues important in kidney function [2]. Whereas previously it was thought that sncRNAs merely reflect degradation by-products, it has now been shown that they exert important functions in cell-to-cell communication, the regulation of gene expression, splicing and ribosomal RNA maturation [2].

Although a substantial number of studies have examined circulating sncRNAs associated with kidney function in persons with diabetes, most studies are relatively small (n<100) and focused on microRNAs. From these studies a large number of miRNAs have emerged that are putatively associated with kidney function. However, probably due to small sample sizes, heterogeneity in endpoints and methodology used there is limited consistency between studies as shown in various meta-analyses [3–5].

To the best of our knowledge there are currently no studies that have systematically examined the other classes (biotypes) of small ncRNA. We thus go beyond the current state-of-the-art by not only investigating microRNAs but also other sncRNAs classes, including besides miRNAs, piRNAs, tRNA fragments (tRFs), lncRNA fragments, Y-RNAs and snoRNAs. For this we applied small RNA sequencing in a large well-defined cohort (n=474) of persons with and without DKD to capture the whole plasma sncRNA transcriptome allowing for a hypothesis-free, comprehensive analysis of the role of sncRNAs in diabetic kidney disease [6].

## Materials and Methods

A schematic overview of the study design can be found in Supplemental Figure 1.

### Study sample

For this project we used data from the Hoorn Diabetes Care System cohort (DCS), a large prospective study comprised of 14,000+ individuals with type 2 diabetes who receive structed diabetes care [7]. Participants visit the Hoorn diabetes research center annually for diabetes care including routine biochemistry, medication review and clinical care. Patients were also invited to participate in DCS biobanks in which we combine the annual phenotypic data with biobanking of liquid biopsies which includes serum, plasma and urine. Blood samples were collected during two phases of biobanking in 2008/2009 and 2012-2014. For the current study we used a subsample from the whole DCS biobank (n=475) using the following inclusion criteria: a plasma sample collected within four years after diagnosis of diabetes and no-insulin use at the time of blood sampling.

All laboratory measurements were on samples taken in a fasted state on the same day as the sample for RNA-seq. Routine anthropometric, biochemistry methods and definitions have been described previously in detail in van der Heijden et al. [7]. Kidney function was assessed based on eGFR (CKD-EPI) and urinary albumin to creatinine ratio (UACR) and included as continuous variables in our analyses. Stages of chronic kidney disease (CKD) were defined according the KDIGO criteria as previously described [8]. In short, this definition uses a combination of eGFR and UACR to categorize persons based on kidney function. Persons in stage 0 do not show signs of kidney dysfunction whereas the stages 1 to 3 have increasing kidney dysfunction as shown in Supplemental Figure 2. Cases were defined as having CKD stage ≥ 1 (i.e. eGFR <60 ml/min/1.73 m^2^ and/or UACR >3 mg/mmol) at the time of blood sampling for sncRNA analysis which resulted in 69 cases.

### Circulating small RNA sequencing

Circulating sncRNAs were isolated from 900ul of citrate plasma using a commercial cell-free RNA isolation kit (Quick-cfRNA Serum & Plasma Kit, Zymo Research, Irvine, CA, USA). After quality control 475 samples were submitted for RNA sequencing. Library preps were constructed using the NEBNext® Multiplex Small RNA Library Prep Set for Illumina (New England Biolabs, Ipswich, MA, USA). A Pippin Prep instrument (Sage Science, Beverly, MA,USA) was used for size selection to further enrich for sncRNAs with a size of ∼20 to 100 nucleotides. Sequencing was performed using the Illumina NextSeq500 sequencer with v2.5 sequencing reagent kits and 35 cycle paired-end reads (PE35). On average 11.3±2.8 million reads per sample were generated. The exceRpt pipeline [9], developed by the NIH Extracellular RNA Communication Consortium (ERCC), was used to map the obtained reads, for quality control, to process the data and generate RNA abundance estimates for miRNA, piRNA, lncRNA, tRNA fragments, Y_RNA (fragments), snRNA, snoRNA, scaRNA and various other sncRNA species. Using respectively miRBase v22 [10], piRNABank v1 [11], Gencode (version 38) [12], circBase [13] and GtRNAdb [14]. One sample did not pass the QC thresholds.

### Taqman validation of the RNA-seq results

We used Taqman qPCR assays (ThermoFisher Scientific, NL) to verify the validity and reproducibility of the small RNA-seq results. First cDNA was generated from 5ng of RNA isolated from plasma using the TaqMan™ Advanced miRNA cDNA Synthesis Kit (ThermoFisher Scientific, NL) according to the instructions of the manufacturer. We chose four miRNA assays with a range from low to relative high expression in our study sample (miR-542-3p, miR-323b-3p, miR-186-5p and miR-423-5p) which were measured in 12 randomly selected plasma RNA samples for which sncRNA-seq data were available. Clinical characteristics are shown in Supplemental Table 1. The Taqman qPCR assays were run according to the manufacturer’s instructions using TaqMan Fast Advanced Master mix and TaqMan Advanced miRNA Assays (ThermoFisher Scientific, NL). miR-191-5p expression was used as the reference (housekeeping gene) in both the Taqman and small RNA-seq datasets as suggested by the manufacturer [15].

### Expression patterns of small RNAs in tissues

Publicly available raw sncRNA-seq data for various metabolic tissues were downloaded from the Gene Expression Omnibus (GEO) [16]. The tissues for which sncRNA-seq data was used included kidney, thyroid, brain, liver, muscle, heart, colon, pancreas, subcutaneous white adipose tissue and whole blood and urine. Accession numbers are given in Supplemental Table 2. Data were processed using the exceRpt pipeline as described above. The tissue data were used to estimate the abundance of the sncRNAs in these tissues. Data was z-scaled in figures.

### Plasma proteomics

Plasma proteomic data was available for 589 participants of the Hoorn DCS study and generated using the SomaLogic SOMAscan platform (Boulder, CO) as previously described [17]. After quality control 1195 protein levels were available for analysis. For 199 subjects we also had the corresponding sncRNA-seq data from the same sampling date. Clinical characteristics of the participants in this study are shown in Supplemental Table 1.

### Statistical analysis

The R-package edgeR (v3.42.4) was used to identify differentially expressed sncRNAs [18]. A negative binomial generalized log-linear model was applied using edgeR’s function *estimateGLMRobustDisp*. For the determination of differential expression and p-values we used the quasi-likelihood (QL) F-test using edgeR’s *glmQLFTest* function according to the recommendations of the developers. The base model was unadjusted. A fully adjusted model was adjusted for BMI, diabetes duration, HbA1c, HDL, SBP and smoking unless otherwise stated. Age and sex were excluded from the model as these are part of the definition of eGFR. UACR was log10-transformed before analysis. In sensitivity analyses, we additionally included use of antihypertensive medication and or eGFR or UACR. The correlation between the sncRNA transcriptome and plasma proteome was tested using a linear model with the log transformed proteomic measurements as outcome and the sncRNA transcriptome as predictors with adjustment for age, BMI, sex and technical covariates. Associations between the proteome and eGFR or UACR were tested using the same model. The Benjamini-Hochberg procedure was used to adjust for multiple hypothesis testing and an FDR below 0.05 was considered significant.

## Results

### Clinical characteristics

The clinical characteristics of our study group are shown in Table 1. The characteristics resemble those of a typical T2D population with slightly more males than females, a mean age-at-diagnosis of 59.3 ± 9.0 years, BMI of 30.7 ± 6.0 kg/m2 and an average duration of diabetes at time of blood sampling of 2.2 ± 1.1 years. Glycemic control was good with an average HbA1c of 46 (42, 51) mmol/mol. Prevalence of DKD at the time of blood sampling was 15%.

**Table 1.** Clinical characteristics of the study population.

|  | all | No DKD | DKD |
| --- | --- | --- | --- |
| n | 475 | 406 | 69 |
| Gender (%Female) | 45 | 46 | 42 |
| Age (years) | 61.4 ± 9.1 | 61.0 ± 8.8 | 63.8 ± 10.1* |
| BMI (Kg/m<sup>2</sup>) | 30.7 ± 6.0 | 30.3 ± 5.6 | 32.7 ± 7.3* |
| Age-at-Diagnosis (years) | 59.3 ± 9.0 | 58.8 ± 8.7 | 61.8 ± 9.8* |
| Diabetes duration (years) | 2.2 ± 1.1 | 2.2 ± 1.1 | 2.0 ± 1.1 |
| Fasting glucose (mmol/L) | 8.0 ± 1.6 | 8.0 ± 1.7 | 8.1 ± 1.4 |
| HbA1c (mmol/mol) | 46 (42, 51) | 46 (42, 51) | 46 (43, 54) |
| HDL (mmol/L) | 1.19 ± 0.32 | 1.20 ± 0.33 | 1.11 ± 0.25 |
| eGFR (ml/min/1.73 m<sup>2</sup>) | 84 ± 16 | 87 ± 13 | 70 ± 22* |
| eGFR ≤ 60 (%) | 34 (7) | 0 | 34 (50) |
| Urine Albumin-to-Creatinine ratio (mg/mmol) | 0.48 (0.20, 1.00) | 0.41 (0.18, 0.79) | 3.99 (0.79, 10.5)* |
| Albuminuria (%) | 43 (9) | 0 | 43 (62) |
| Chronic Kidney Disease (%) | 69 (15) | 0 | 69 (100) |
| Diastolic blood pressure (mm Hg) | 78 ± 9 | 78 ± 9 | 79 ± 10 |
| Systolic blood pressure (mm Hg) | 139 ± 18 | 139 ± 18 | 142 ± 20 |
| Smoking (current/ever/never) | 92/130/250 | 76/107/220 | 16/23/30 |
| Medication use |  |  |  |
| Metformin (%) | 340 (72) | 287 (71) | 53 (77) |
| SU (%) | 130 (27) | 105 (26) | 25 (36) |
| Other glucose-lowering (%) | 13 (3) | 12 (3) | 1 (1) |
| Antihypertensive medication (%) | 319 (67) | 260 (64) | 59 (86)* |
| Lipid lowering medication (%) | 320 (67) | 271 (67) | 49 (71) |

Data are mean ± SD, n (%) or median (IQR). All measures taken at the time of blood sampling.
\* P≤0.05 versus those without DKD.

### sncRNA biotype abundance in our samples

After quality control, 938 sncRNAs were expressed above the threshold of a mean ≥5 copies in our data, which included miRNAs, piRNA, Y_RNA fragments, lncRNA fragments, tRNA_fragments, snoRNAs, snRNAs and various other sncRNA species (Table 2). miRNAs, Y-RNAs and tRFs were the most abundant sncRNAs biotypes comprising respectively 57.6, 34.9 and 4.8 percent of all reads. Other sncRNA biotypes were much less abundant (≤1.4%). There were no significant differences in the total number of reads per biotype between cases and controls (all P>0.4).

**Table 2.** Frequency of different types of sncRNAs associated with the three endpoints.

|  | Total # of<br>sncRNA<br>detected |  |  | DKD |  |  |  | eGFR |  |  |  | UACR |  |  |
| --- | --- | --- | --- | --- | --- | --- | --- | --- | --- | --- | --- | --- | --- | --- |
|  | n | % |  | n | % | p* |  | n | % | p* |  | n | % | p* |
| Total # sncRNAs | **938** | **100** |  | **7** |  |  |  | **107** |  |  |  | **220** |  |  |
| miRNA | 602 | 64 |  | 1 | 14 | 0.011 |  | 63 | 59 | 0.29 |  | 93 | 42 | 4.5x10<sup>-9</sup> |
| snoRNA | 96 | 10 |  | 5 | 71 | 2.2x10<sup>-4</sup> |  | 14 | 13 | 0.40 |  | 67 | 31 | 8.4x10<sup>-13</sup> |
| lncRNA | 93 | 10 |  |  |  |  |  | 13 | 12 | 0.50 |  | 20 | 9 | 0.80 |
| piRNA | 45 | 5 |  | 1 | 14 | 0.30 |  | 11 | 10 | 0.037 |  | 23 | 11 | 4.0x10<sup>-3</sup> |
| Y_RNA fragments | 33 | 4 |  |  |  |  |  |  |  |  |  | 1 | 0.5 | 0.013 |
| others | 26 | 3 |  |  |  |  |  |  |  |  |  | 6 | 3 | 1.00 |
| tRNA fragments | 24 | 3 |  |  |  |  |  | 4 | 4 | 0.52 |  | 1 | 0.5 | 0.067 |
| snRNA | 12 | 1 |  |  |  |  |  | 1 | 1 | 1.00 |  | 6 | 3 | 0.13 |
| scaRNA | 7 | 1 |  |  |  |  |  | 1 | 1 | 0.58 |  | 3 | 1 | 0.41 |

\* P-value for comparison against the distribution of all sncRNAs detected.

### Taqman validation shows good reproducibility

We next sought to verify validity and reproducibility of the sncRNA-seq. Taqman qPCR assays for sncRNAs with high, medium and low expression were used and these showed a good correlation with the RNA-seq results (r^2^= 0.86, Supplemental Figure 3).

### sncRNAs associate with DKD, eGFR and UACR

Seven sncRNAs were significantly associated with prevalent DKD in the unadjusted model (P_FDR_≤0.05, Table 3, Figure 1A). All identified sncRNAs were downregulated in DKD cases. The top hit was snoRNA U8 (log2 fold change=−0,859, P_FDR_=1.90×10^−3^, Figure 1B). Five of these sncRNAs also showed a significant trend for lower levels in those with more advanced stages of CKD (P_FDR_≤0.05, Supplemental Figure 4).

**Figure 1.**
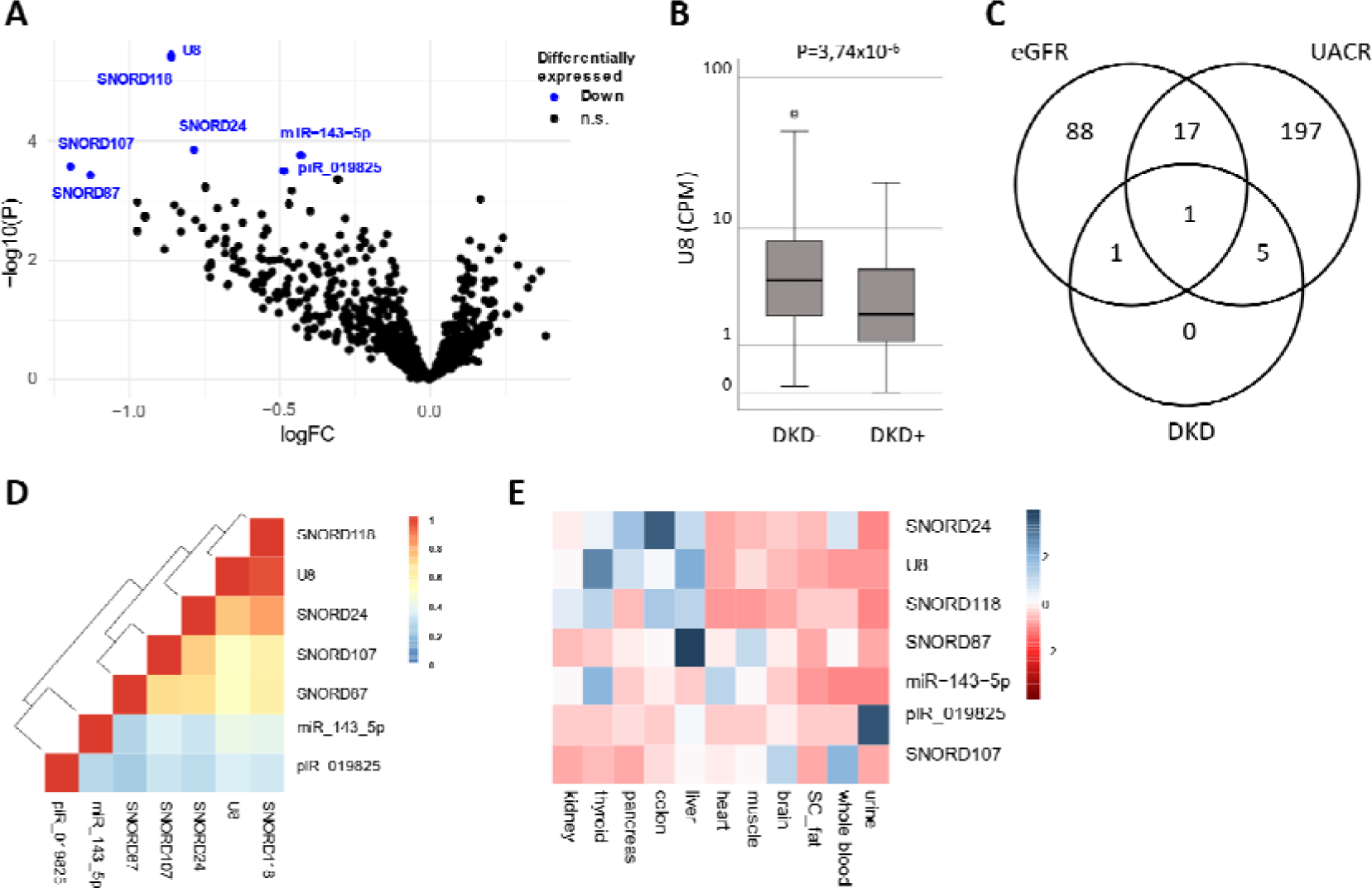
sncRNAs significantly associated with DKD. Panel A. Volcano plot showing the association between sncRNAs and DKD in the base model. sncRNAs in blue are significantly lower is cases with DKD after multiple hypothesis testing (FDR≤0.05). logFC is the Log2 of the fold change between cases and controls. Panel B. U8 levels in those with or without DKD. CPM = counts per million reads. Panel C. Overlap between sncRNAs significantly associated with DKD, eGFR or UACR in the base model. Panel D. Pairwise correlations for the seven sncRNAs associated with DKD. Colors represent Spearman’s rho. Panel E. Expression pattern for sncRNAs associated with DKD in publicly available datasets for 11 tissues of interest. Colors represent the Z-scaled expression of a sncRNA where blue indicates relative high expression and red relative low expression.

**Table 3.** Overview of seven sncRNAs associated with prevalent DKD.

| sncRNA | Prevalent DKD |  | eGFR |  | UACR |  |
| --- | --- | --- | --- | --- | --- | --- |
|  | LogFC | P<sub>FDR</sub> | logFC | P<sub>FDR</sub> | logFC | P<sub>FDR</sub> |
| U8 | -0,859 | 1,90x10<sup>-03</sup> | 0,012 | 2,79x10<sup>-02</sup> | -0,724 | 6,74x10<sup>-03</sup> |
| SNORD118 | -0,861 | 1,90x10<sup>-03</sup> | 0,009 | 8,94x10<sup>-02</sup> | -0,858 | 1,73x10<sup>-03</sup> |
| miR-143-5p | -0,429 | 4,01x10<sup>-02</sup> | 0,000 | 9,86x10<sup>-01</sup> | -0,376 | 3,30x10<sup>-02</sup> |
| SNORD24 | -0,785 | 4,01x10<sup>-02</sup> | 0,011 | 6,42x10<sup>-02</sup> | -0,919 | 1,73x10<sup>-03</sup> |
| piR_019825\|DQ597218 | -0,485 | 4,91x10<sup>-02</sup> | 0,012 | 3,04x10<sup>-03</sup> | -0,129 | 5,71x10<sup>-01</sup> |
| SNORD107 | -1,194 | 4,91x10<sup>-02</sup> | 0,013 | 1,43x10<sup>-01</sup> | -2,214 | 1,58x10<sup>-05</sup> |
| SNORD87 | -1,129 | 4,91x10<sup>-02</sup> | 0,012 | 2,17x10<sup>-01</sup> | -2,253 | 1,18x10<sup>-05</sup> |

LogFC is the log2 of the fold change. P<sub>FDR</sub> represents the P value after correction for multiple hypothesis testing using the Benjamini-Hochberg procedure.

Respectively, 107 and 220 sncRNAs were significantly associated with eGFR and UACR, which are used for the DKD classification (Table 3, Supplemental Table 3, Supplemental Figures 5 and 6). The overlap between the sncRNAs associated with DKD and its components eGFR and UACR is shown in figure 1C and Table 3. snoRNA U8 overlapped between all three endpoints used and piR_019825 overlapped between eGFR and DKD. SNORD118, SNORD24, SNORD107, SNORD87 and miR-143-5p all overlapped between UACR and DKD.

In the full model respectively 7, 153 and 205 sncRNAs remained significantly associated with DKD, eGFR and UACR (Supplemental Table 4, Supplemental Figure 7; 57%; 95% and 84% overlap with the unadjusted model). U8, SNORD118, piR_019825 and miR-143-5p were significant in both the base and full models. In a sensitivity analysis, additional adjustment for use of antihypertensive medication and baseline eGFR or UACR (only for eGFR and UACR) did not have a major impact on the effect sizes but reduced the number of significant associations (r^2^>=0.92, Supplemental Figure 8).

### Certain types of sncRNAs are enriched in DKD

Comparing the frequency of different types of sncRNAs identified showed that while snoRNAs represent 10% of the total small RNA transcriptome in our RNA-seq study they are the most abundant class associated with prevalent DKD (71%, OR=21.9, P=2.4×10^−4^, Table 2). Enrichment for snoRNAs is also seen for UACR (OR=3.8, p=8.4×10^−13^). Associations with miRNAs on the other hand are less frequent whilst piRNAs also seem enriched (Table 2). A similar pattern is again seen for UACR and to a lesser extend for eGFR. In the full model a largely similar picture is observed (Supplemental Table 5).

The pairwise correlation in expression between the seven sncRNAs associated with diabetic DKD was very modest (Figure 1D). The only exceptions were U8 and SNORD118, which is likely due to the fact that they are encoded in the same host gene (TMEM107) on chromosome 17. Results of these analyses for eGFR and UACR are shown in Supplemental Figure 9.

### sncRNAs are tissue-enriched and correlate with plasma protein levels

In a next step, we examined the tissue expression patterns of the identified sncRNAs. We used publicly available expression data in 11 tissues or body fluids (Fig. 1E). This illustrated that some sncRNAs show tissue-enriched expression patterns. For the seven sncRNAs associated with DKD none had its highest expression in kidney tissue. piRNA-019825 was the only sncRNA that showed high expression in urine. Similar observations were made for eGFR and UACR associated sncRNAs (Supplemental Fig. 10).

Finally, since the function of most of the sncRNAs we found in our study is unknown we compared the expression levels of the sncRNAs with expression of 1195 proteins measured in plasma from the same date to identify potential targets or (co-)regulated proteins or pathways. As shown in Supplemental Table 6 we found for the seven sncRNAs 31 FDR significant pairwise correlations involving 24 proteins. The strongest association was observed between piR-019825 and TNF sR-1 (TNFRSF1A, Beta=−0.080 (−0.112 - −0.040), P_FDR_=3.04×10^−2^). piRNA-019825 also showed the largest number of correlations (n=11, Figure 2A). Interestingly, among the proteins associated with piR-019825 was Cystatin C, a known marker of kidney function. Next we assessed whether the 24 proteins were also significantly associated with eGFR or UACR (FDR≤0.05). Six were associated with eGFR and none with UACR. Four out of the six proteins that were negatively associated with piR-019825 expression were also negatively associated with eGFR (all P_FDR_≤3.78×10^−11^) (Figure 2A, Supplemental Table 7). Two proteins were associated with miR-143-5p expression and another eleven proteins were associated with snoRNA expression. Five proteins were correlated with two or more snoRNAs (NAMPT, UFM1, PDGFB, CCL27 and CCL28) and two were also significantly associated with eGFR (CCL27, RARRES2, Figure 2B). Pairwise correlations between the piR-019825 or snoRNA associated proteins is shown in figures 2C, D

**Figure 2.**
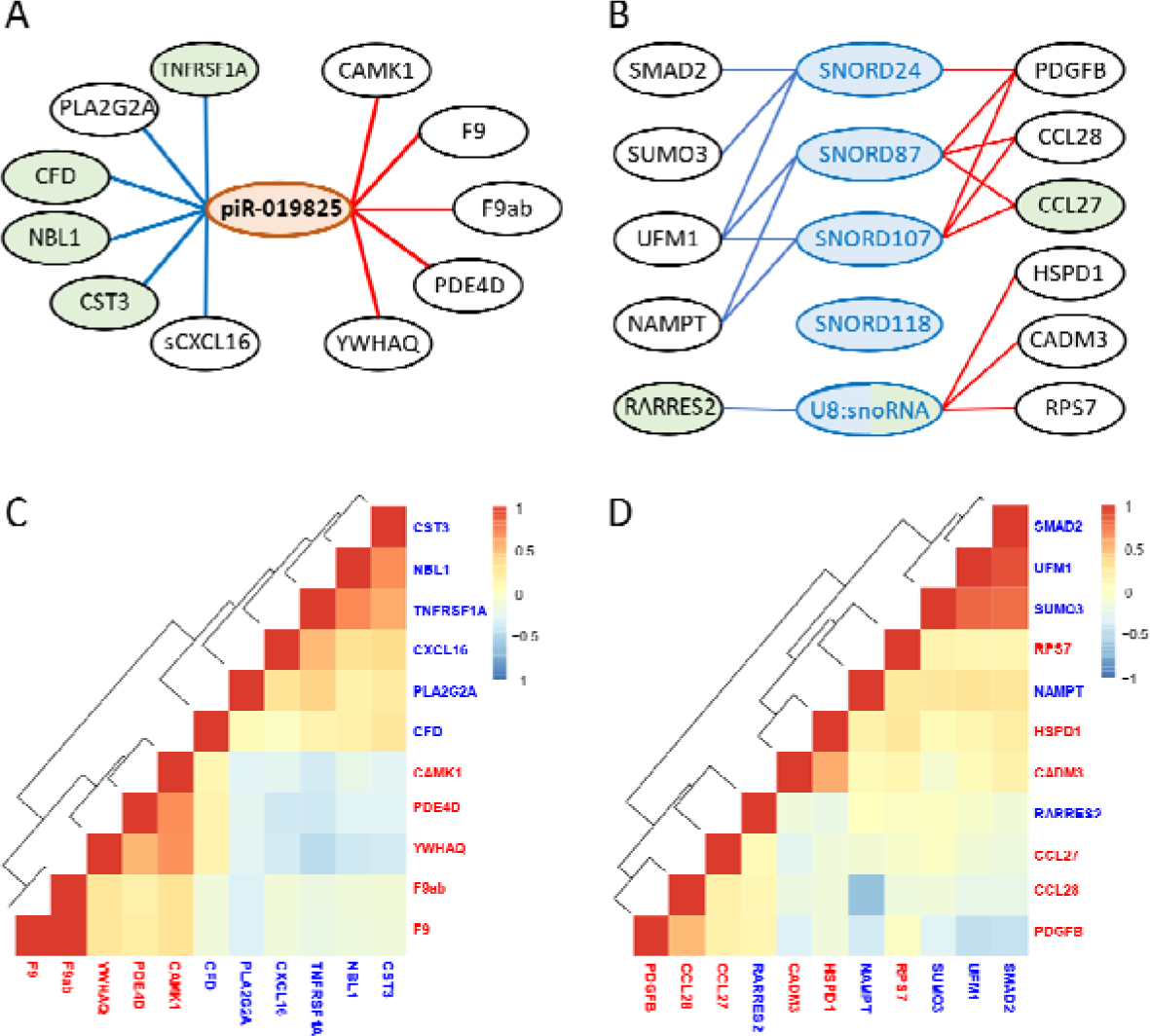
sncRNAs associated proteins. Overview of the proteins that associate with sncRNA expression levels in plasma from persons with T2D. sncRNAs and proteins in green are also significantly associated with eGFR and those in blue are associated with UACR (P_FDR_≤0.05). Blue line and text indicate a negative association and a red line and text indicates a positive association. Panel A. piR-019825 associated proteins (P_FDR_≤0.05). Panel B. snoRNA associated proteins (P_FDR_≤0.05). Panel C. Pairwise correlations between the 11 piR-019825 associated proteins. Panel D. Pairwise correlations between the 11 snoRNA associated proteins.

## Discussion

In this study we show that circulating sncRNAs are associated with prevalent DKD in people with type 2 diabetes. Importantly these sncRNAs do not only represent miRNAs but also other classes of small non-coding RNAs. In addition, these sncRNAs correlate with proteins that function in pathways known to be involved in DKD/kidney function.

### Associations between kidney function and sncRNAs

#### miRNAs

MicroRNAs are involved in the post-transcriptional regulation of gene expression and are known to be involved in crosstalk between (distant) cells and tissues [2]. As such, there is an ever growing interest to study their role in disease development and as biomarkers of disease and disease progression.

In our study one miRNA, miR-143-5p, was associated with both DKD and UACR. Levels of this miRNA are lower in both DKD and persons with high UACR. miR-143-5p remained significant in the full models, suggesting it might have biomarker potential. Previous in vitro studies showed that this miRNA is associated with vascular pathology [19], an impaired glomerular filtration barrier [20] and with mesangial cell proliferation and fibrosis [21, 22]. miR-143-5p plasma levels associate with two proteins but neither of those have a clear relationship with kidney function nor were they predicted targets for miR-143-5p (miRDB, miRTarBase, MiRBase). In the full model three additional microRNAs were significant, miR-455-3p, miR-122-3p and miR-193b-3p. Previously it was shown that miR-455-3p was down regulated in human mesangial cells (HMC) and human proximal tubule epithelial cells (HK-2) under DKD stimulating conditions [23]. Interestingly, it was also shown that overexpression of this miR could alleviate the pathological changes such as renal fibrosis in a rat DN model [23]. In a mouse mesangial cell line miR-455-3p improved cell proliferation, inflammation and ECM accumulation induced by high glucose treatment [24]. Involvement of this miR in fibrosis in other tissues like liver and lung has also been reported. A previous report indicated that miR-122, a liver specific miR associated with liver injury, was reduced in patients with end stage renal disease but not in CKD when compared to healthy controls [25]. miR-193b-3p was identified as a potential biomarker for pre-diabetes and also for CKD after nephrectomy for renal cell carcinoma [26, 27].

In addition to the association with DKD we also found many miRNAs associated with eGFR and or UACR. A large number of which remained significant in the full models. Interestingly, of the 154 miRNAs found in the base models for eGFR and UACR only two of them were associated with both phenotypes suggesting that the associated miRNAs are specific to either defect in kidney function but not both.

#### piRNAs

PIWI-interacting RNAs are involved in silencing of transposons but they can also be involved in the regulation of gene expression in a miRNA-like manner [28]. One piRNA, piRNA-019825 (DQ597218), was significantly associated with DKD. Previously, it was shown that this piRNA is one of the most abundant piRNA species in urine and plasma [29]. In our proteomics analysis to identify potential targets or pathways we found eleven proteins whose expression was correlated with the levels of piR-019825 including cystatin C, a known marker for kidney function. Four of the negatively associated proteins are also strongly associated with eGFR. This is in line with a putative miRNA like function for piR-019825 in which lower piRNA expression would result in higher protein levels, lower eGFR and increased risk of DKD. This is further corroborated by the fact that increased levels of two of these proteins were previously found to increase risk of CKD or end stage renal disease (ESRD) in prospective studies in persons with diabetes (TNF sR-I (TNFRSF1A), NBL1 (DAN) [30–33]. Whereas the other negatively associated proteins we found associated with this piRNA (CST3, sCXCL16, CFD, PLA2G2A) have almost all also been implicated previously in kidney dysfunction [34–36]. Interestingly, we also find various other piRNAs associated with either eGFR and or UACR (10-14%). Most of which remained significant in the full models. Suggesting that they may be useful early markers of DKD. Of note, eGFR-associated piRNAs often show their highest expression in urine, whereas UACR associated piRNAs show low expression in urine, but often high levels in tissues like pancreas, liver and colon.

#### snoRNAs

Small Nucleolar RNAs (snoRNA) are predominantly localized to the nucleolus where they have a function in rRNA biogenesis, structure and activity [37]. There are two major types of snoRNAs containing either box C/D or box H/ACA motifs. In our study we only detected box C/D snoRNAs associated with DKD. Box C/D snoRNAs are involved in 2’-O-methylation of specific sequences in target RNA sequences, mainly rRNA, thereby affecting RNA structure, stability and interactions. In addition, other functions have been ascribed to the snoRNAs such as mRNA splicing and editing. SnoRNA derived fragments have also been described and it has been speculated that they exert piRNA or miRNA like functions [38].

Although snoRNAs represent only ten percent of the total circulating sncRNA transcriptome in our study they represent seventy-one percent of the sncRNAs found associated with DKD suggesting that snoRNAs may function as early biomarkers of DKD. SnoRNA U8 was associated with all three phenotypes but has not been linked to kidney function previously. SnoRNAs SNORD118, SNORD24, SNORD107 and SNORD87 where associated with DKD and UACR but not eGFR. SnoRNAs U8 and SNORD118 both remained significant in the full models. SNORD87 and SNORD107 associate with the same set of proteins, including UFM1, NAMPT, PDGFB, and the two chemokines CCL27 and CCL28, which bind to the same receptor (CCR10). A recent report by Chen et al. showed that post-transcriptional stabilization of the CCL28 mRNA by N6-methyladenosine (m6A) modification increases protein levels of this chemokine and reduces kidney damage after ischemia reperfusion injury [39]. Given the known role of snoRNAs in post-transcriptional RNA modification and stabilization via 2’-O-methylation this might provide a putative mechanism and explanation for the observed snoRNA-protein associations. The other proteins associated with these snoRNAs (UFM1, NAMPT, PDGFB) have also been implicated in kidney function previously [40–43]. Further studies are needed to investigate the role of these snoRNAs in DKD. In addition to the snoRNAs described above various other snoRNAs are associated with eGFR and UACR (13-31% of total number of significant sncRNA). Almost all remained significant in the full models. At present there are no other studies that have investigated the role of snoRNAs in diabetic DKD.

### Members of other sncRNA classes

Although other classes of sncRNAs, such as Y_RNA derived fragments, tRNA fragments (tRF), lncRNA fragments and snRNAs represent roughly twenty percent of all circulating sncRNA detected we did not find any significant associations with DKD. We did find a small number of associations with eGFR and UACR though.

### Strengths and limitations

Our study has several strengths. It is one of the largest circulating sncRNA profiling studies studying the role of circulating sncRNAs in DKD. This improves power and reduces the chance of false positives. In addition, it uses untargeted RNA sequencing allowing not only the study of miRNAs but also several other classes of sncRNA showing that especially these other types of sncRNAs associated with DKD and its components eGFR and UACR. The use of early, mainly mild cases of DKD facilitates the identification of early markers of disease and may help to introduce early interventions aiming to preserve kidney function in those at risk.

A limitation of our study is the use of a total RNA isolation procedure and as such we were not able to investigate if the sncRNAs are carried by extracellular vesicles or are associated with carriers such as HDL and Ago-2 [2]. The use of plasma for RNA isolation is another limitation. Although from a biomarker perspective this is not a limitation, from a functional perspective the role of the sncRNAs we identified needs to be established in the relevant cells and tissues. Another limitation of our study is its cross-sectional, observational design. As such we cannot exclude that (part of) the observed changes and associations are a reflection of the reduced kidney function in the participants.

In conclusion, in this study we have shown that several circulating sncRNAs are associated with DKD. Interestingly, only a relatively small fraction belong to the well-studied microRNAs. This highlights the potential involvement of other classes of sncRNAs, such as snoRNAs and piRNAs, that have so far not been investigated in relation to kidney function before. Future prospective and functional studies are needed to establish the role of these novel circulating sncRNAs in subjects with diabetes and chronic kidney disease.

## Acknowledgements

We would like to thank all staff and participants of the Hoorn Diabetes Care System for their support and participation.

## Author Contributions

LMtH, JWJB and RCS designed the study. JAdK and SJC isolated RNA and prepared the samples for RNAseq. MTB and JWJB were responsible for phenotypic data acquisition and preparation. LMtH, GAB and RCS prepared, analyzed and interpreted the data. LMtH and RCS wrote the first draft of the manuscript. RB contributed to the critical interpretation of the results. LMtH and RCS are the guarantors of this work and, as such, had full access to all the data in the study and take responsibility for the integrity of the data and the accuracy of the data analysis. All authors critically read and revised the manuscript and approved the final version of the manuscript.

## Ethics approval

This study was performed in line with the principles of the Declaration of Helsinki. The study has been approved by the ethics committee of the Amsterdam University Medical Center, location VUmc (approval number 2007/57; NL14692.029.07).

## Consent to participate

All participants provided written informed consent prior to participating in this study.

## Data availability

The datasets generated during and/or analyzed during the current study are not publicly available due restrictions in the informed consent but are available from the corresponding author on reasonable request.

