## Supplementary material for "Small RNA sequencing reveals snoRNAs and piRNA-019825 as novel players in diabetic kidney disease": Sup figures

Supplemental tables see excel file

**Supplemental figure 1. Schematic overview of the study.**

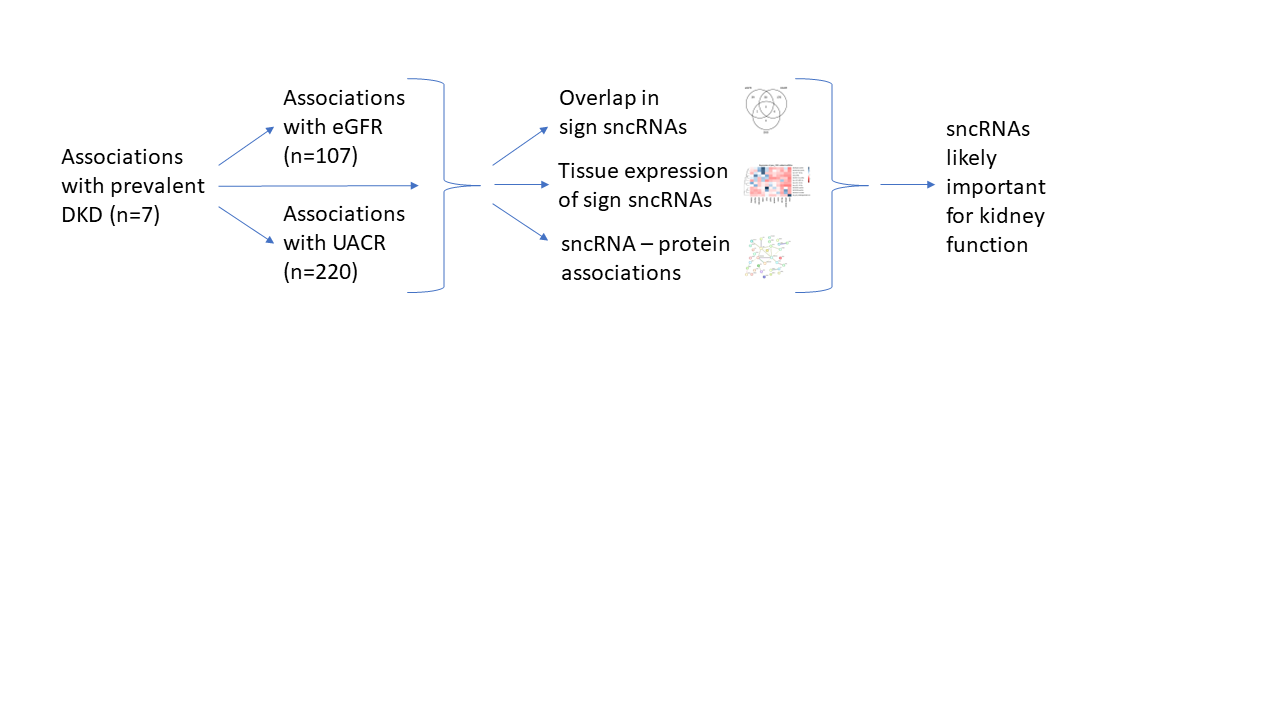

**Supplemental figure 2. Technical validation of the small RNAseq results using Taqman assays.**

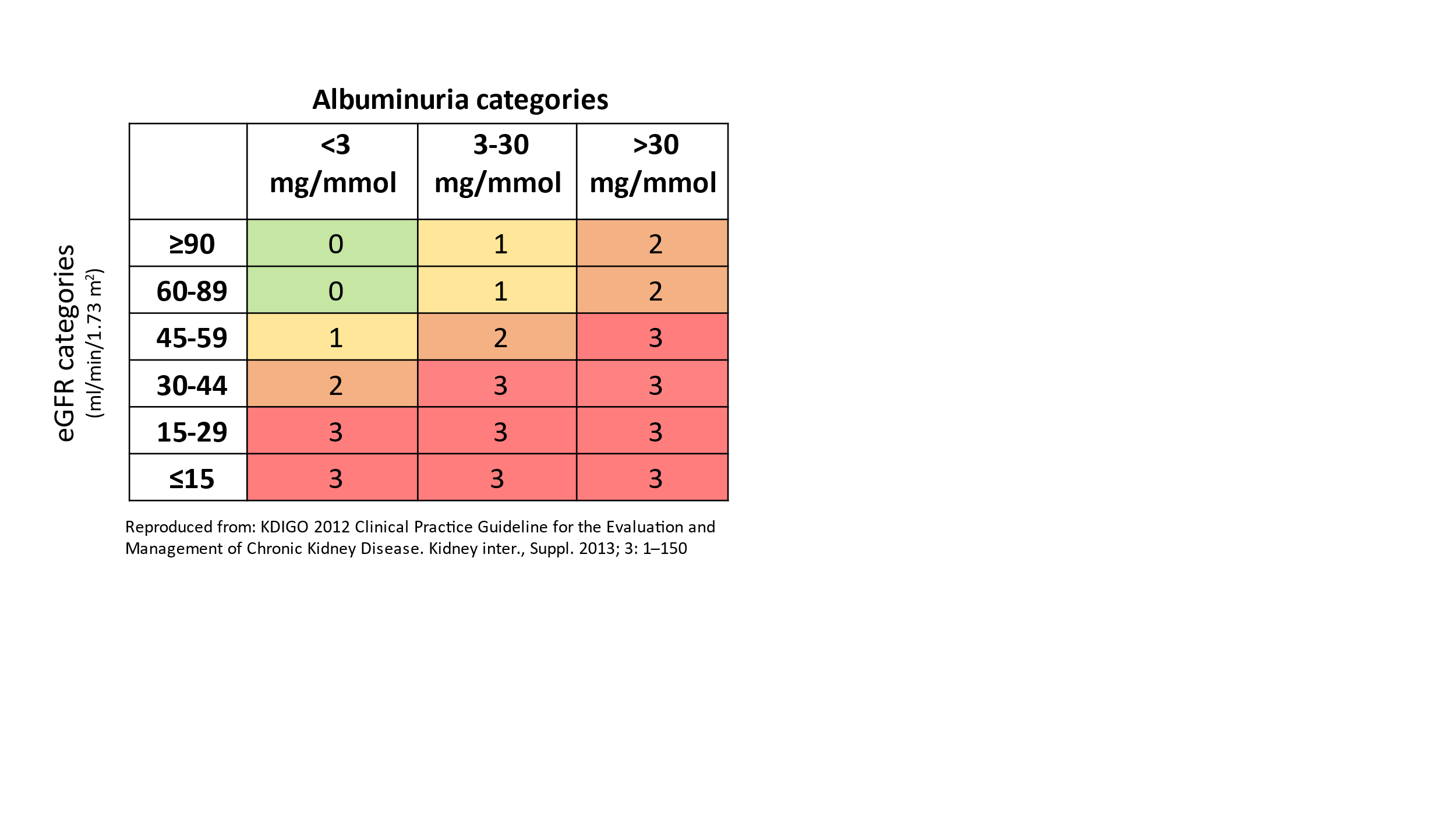

**Supplemental figure 3. Technical validation of the small RNAseq results using Taqman assays.**

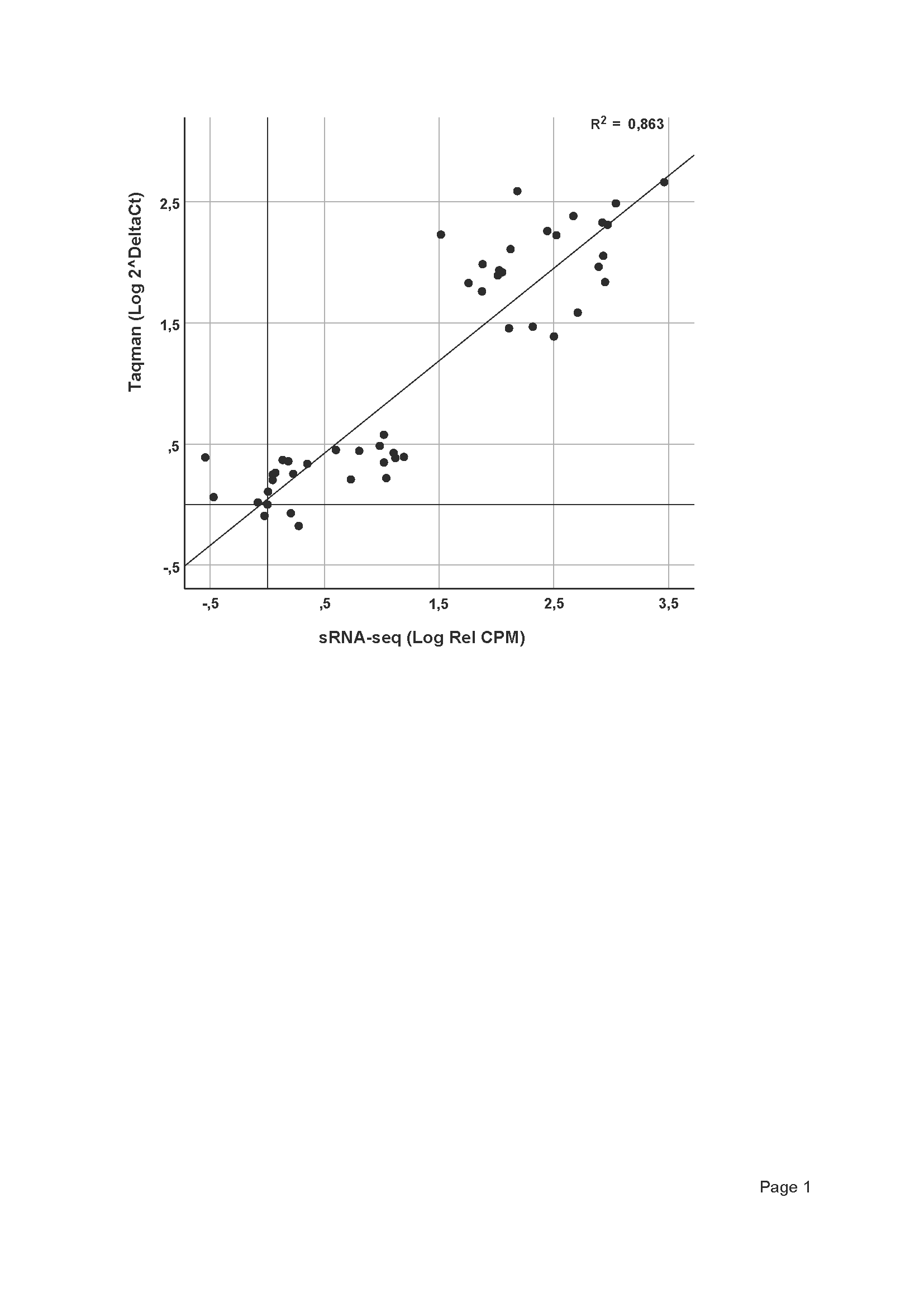

Validation of the sncRNA-seq results using Taqman assays. Data are expressed as log transformed relative expression levels for both Taqman and sncRNA-seq. miR-191-3p expression was used as the reference in both datasets as suggested by the manufacturer (housekeeping gene). For the validation we choose four miRNAs with a range from low to relative high expression (miR-542-3p, miR-323b-3p, miR-186-5p and miR-423-5p) which were measured in 12 plasma samples for which sncRNAs-seq data were available.

**Supplemental figure 4. DKD associated sncRNA levels per CKD-stage**

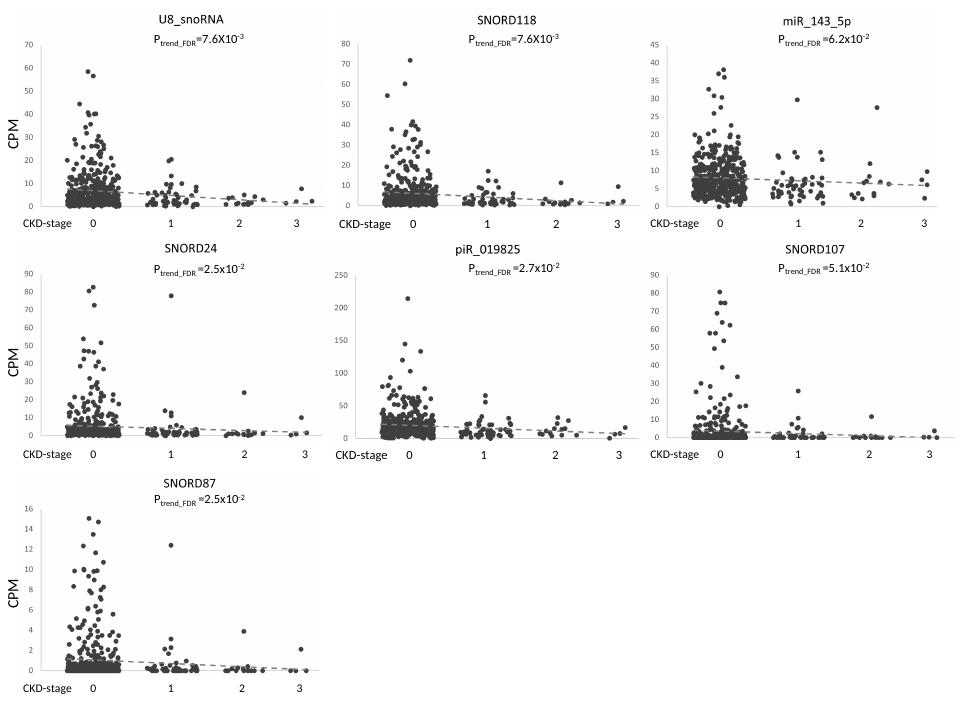

Data represent individual sncRNAs levels per CKD stage. sncRNAs levels were used as outcome, CKD-stage as predictor using the base model. P-values represent FDR corrected P-values.

**Supplemental Figure 5. Volcano plots of the association between eGFR and UACR and small RNAs.**

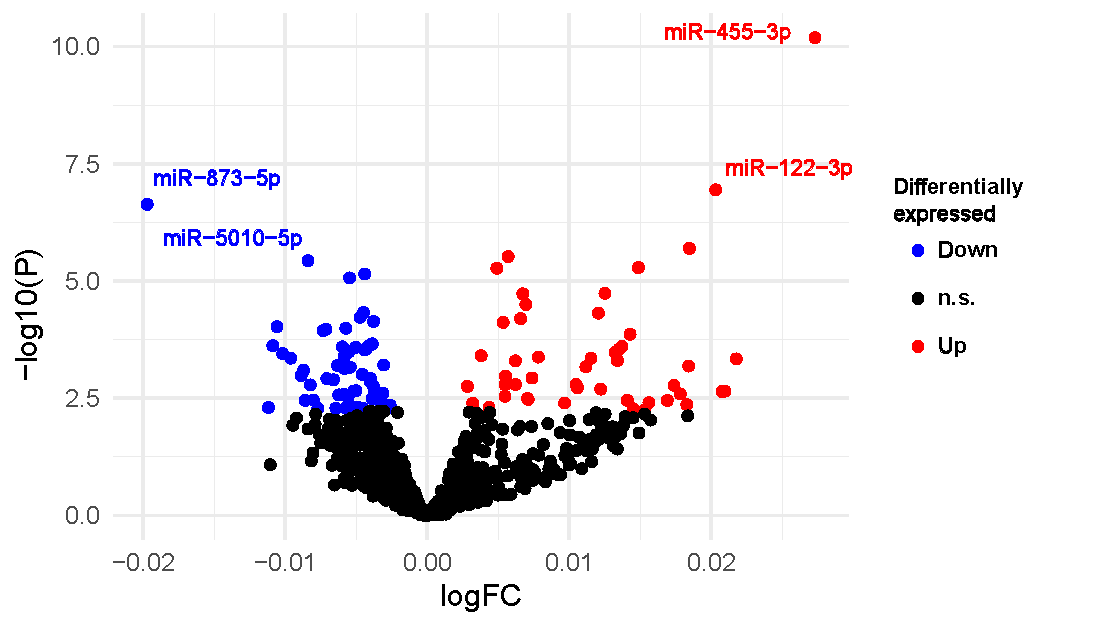

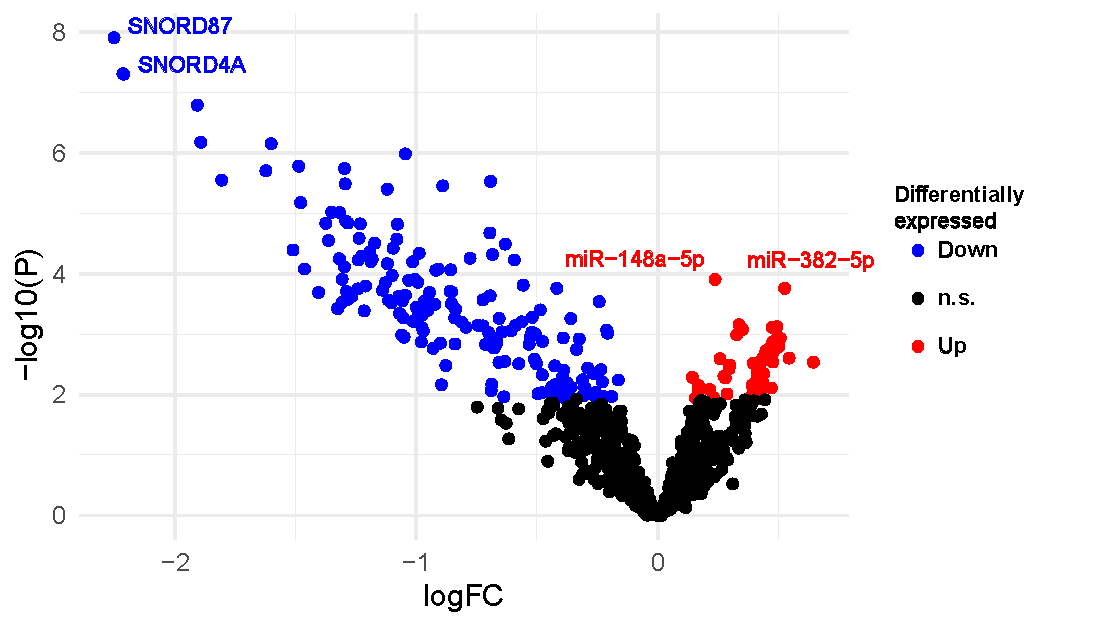

Left panel eGFR and right panel UACR. sncRNAs in blue show a negative association whereas those in red are positively associated after correction for multiple hypothesis testing in the base model (FDR≤0.05). logFC is the Log2 of the fold change between cases and controls.

**Supplemental figure 6. Scatterplots for the four top significant sncRNAs in the base models for eGFR and UACR.**

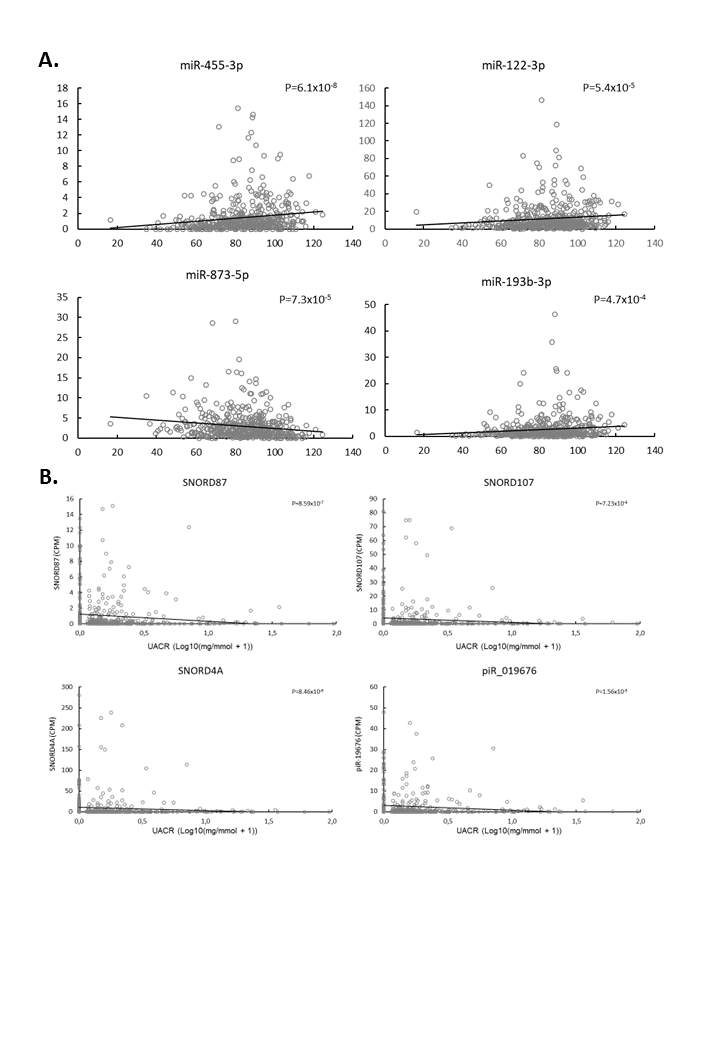

Panel A. eGFR and panel B UACR.

**Supplemental figure 7. Venn diagram showing the overlap in FDR significant sncRNAs for the full model.**

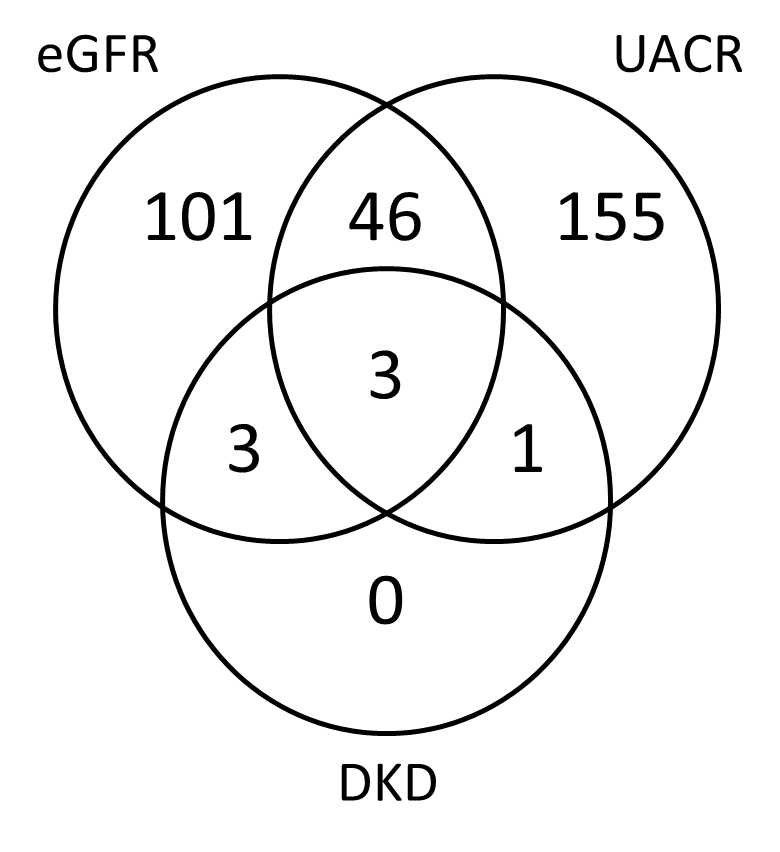

**Supplemental figure 8. Sensitivity analyses including additional adjustments of the full model.**

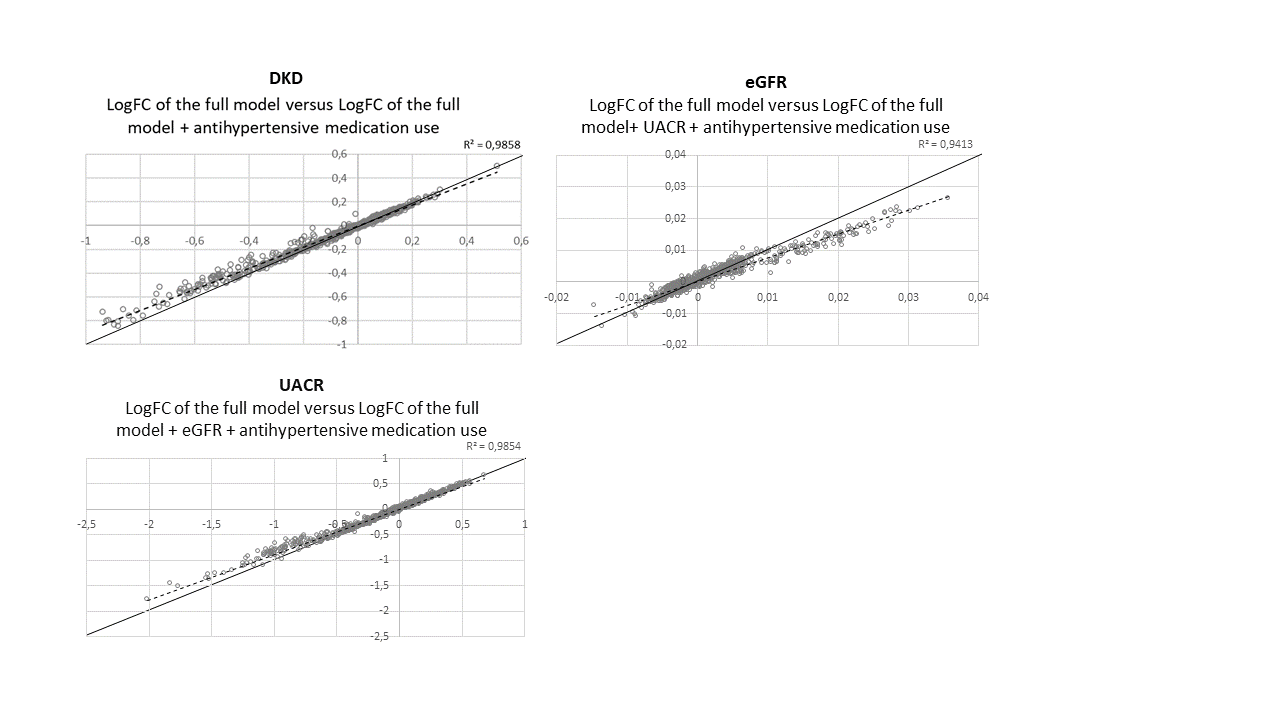

Plots showing the correlation between logFC of the full models versus the logFC of the full model + additional adjustment for the use of antihypertensive medications and eGFR (for the UACR analysis only) or UACR (for the eGFR analysis) only). LogFC of the full models on the x-axis.

**Supplemental figure 9. Pairwise correlations between sncRNAs associated with eGFR or UACR**

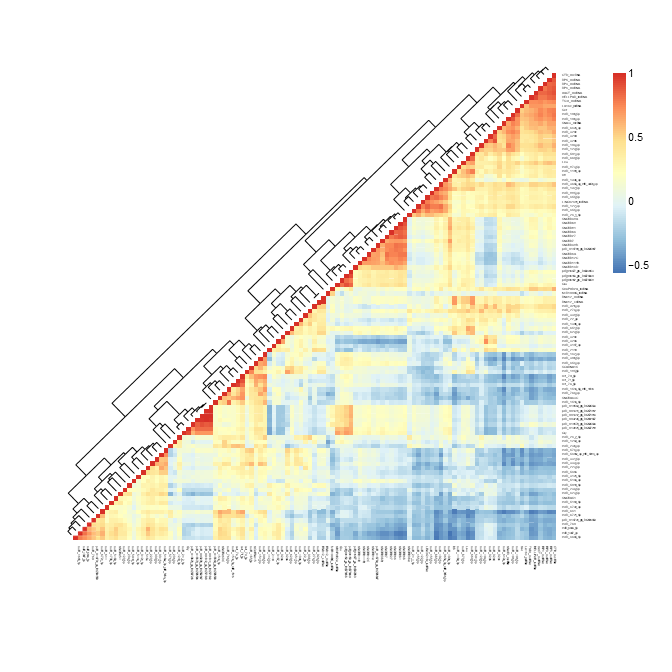

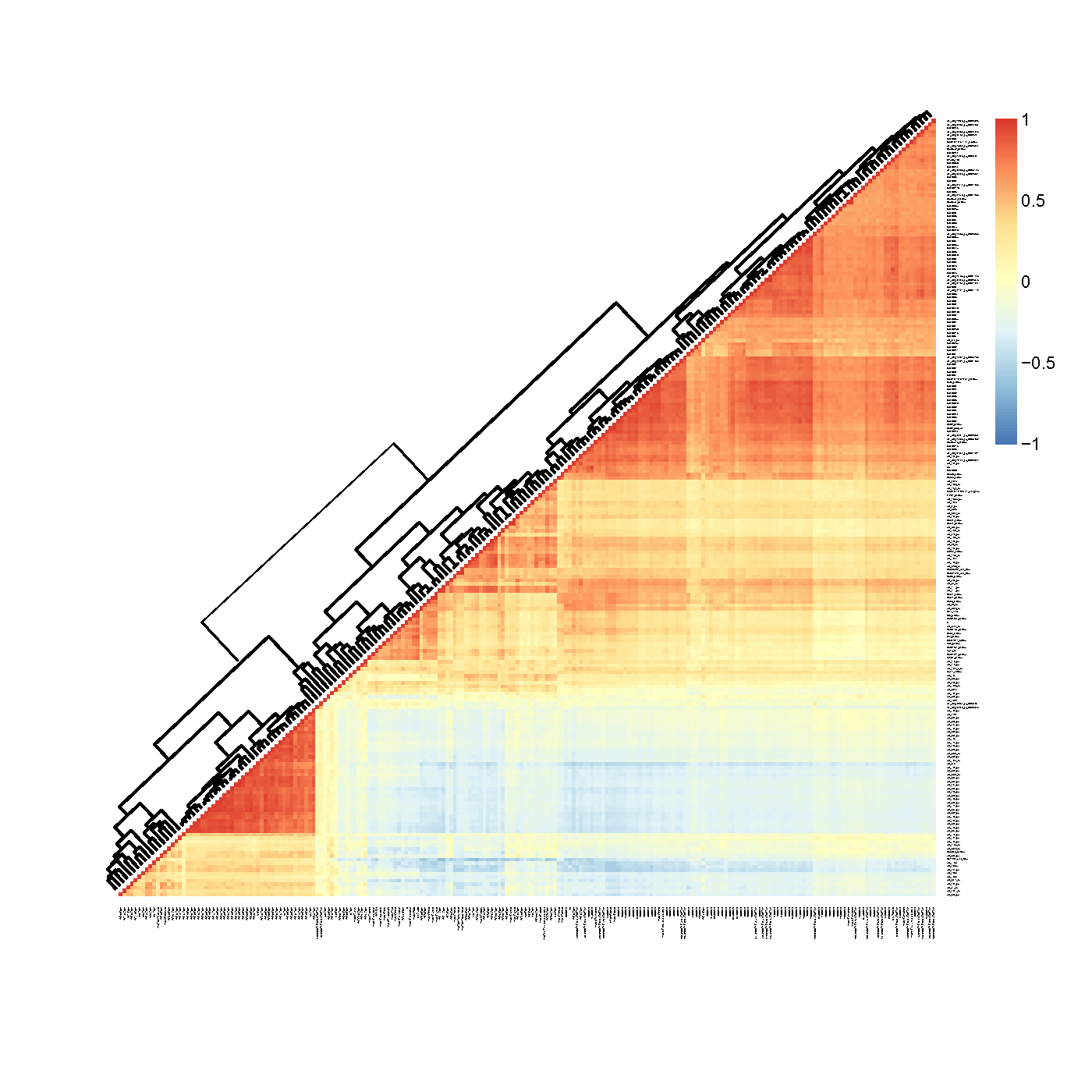

Pairwise correlations for the twelve sncRNA associated with eGFR (left) and UACR (right) in the base model. Colors represent Spearman’s rho.

**Supplemental figure 10** **Expression patterns in various tissues of small ncRNAs associated with eGFR or UACR**

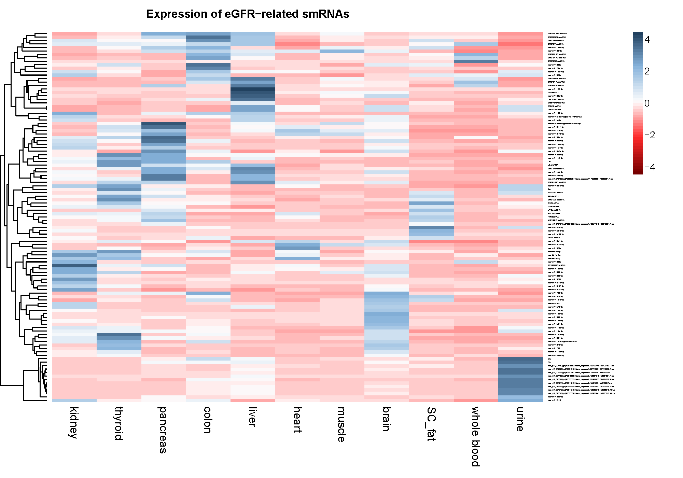

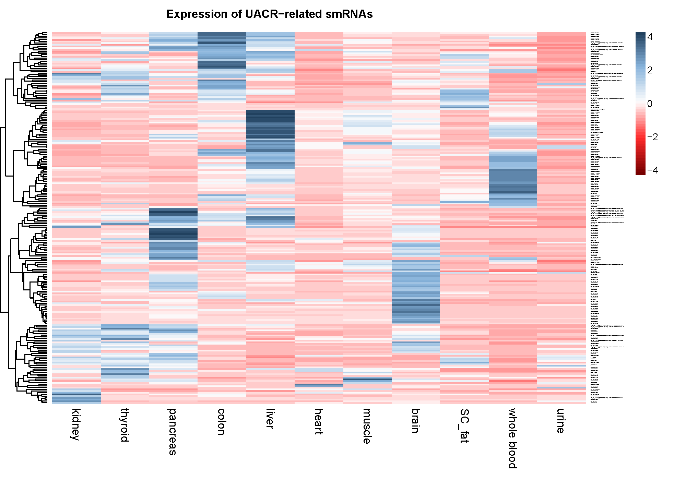

Expression pattern for sncRNAs associated with eGFR or UACR in publicly available datasets for 11 tissues of interest. Colors represent the Z-scaled expression of a sncRNA where blue indicates relative high expression and red relative low expression.
